# AI-Based Coronary Artery Calcification on Non-contrast CT: Performance Across Calcium Scoring, Lung Cancer Screening, and Liver Transplant Candidate Cohorts

**DOI:** 10.64898/2026.05.12.26352904

**Authors:** Kai D. Ludwig, Charles R. Hatt, Lauren Keith, Alexander W. Matyga, Helen S. Te, Luis Landeras, Lydia Chelala, Amit R. Patel, Jonathan H. Chung

## Abstract

**Objective:** Coronary artery calcification (CAC) assessment for cardiovascular risk stratification is traditionally achieved using ECG-gated computed tomography (CT). Automated deep-learning (DL) algorithms may streamline opportunistic CAC detection and scoring, particularly on non-gated CT scans. This study evaluated the performance of a fully automated DL-based CAC scoring algorithm (“DL-CAC”) against expert human scoring.

**Methods:** The algorithm was trained on 1,260 chest CT scans from multiple databases to automatically identify coronary calcium, calculate Agatston scores, and assign a cardiovascular disease (CVD) risk classification. Performance was assessed on a holdout dataset (n=500) comprising ECG-gated calcium scoring CT scans and lung cancer screening non-gated chest CTs as well as in an external, independent CT dataset (n=129) from liver transplant candidates. Agreement with expert scoring was assessed using intraclass correlation coefficient (ICC) for Agatston scores and Cohen’s κ for CVD risk classification.

**Results:** The algorithm demonstrated high agreement with expert scoring in the pooled calcium scoring and lung cancer screening cohorts, with an ICC of 0.947 for Agatston scores and κ of 0.936 for CVD risk classification. For liver transplant candidates, the algorithm exhibited substantial agreement with expert scoring of non-gated CT scans (κ=0.79) and a sensitivity of 90.4% and specificity of 96.4% in high-risk cases.

**Conclusion:** These findings suggest that DL-based CAC scoring on non-gated CT scans may be a feasible alternative to traditional methods and could support opportunistic cardiovascular risk assessment in routine imaging. Further validation is warranted to assess clinical integration in broader practice settings.

## Introduction

Accurate assessment of coronary artery calcification (CAC) is essential for cardiovascular disease (CVD) risk stratification and clinical management in at-risk patients. CAC scoring is a reliable predictor of future CVD events even in asymptomatic populations as it is related to risk of myocardial infarction and sudden death [1, 2]. Traditionally, CAC is quantified using ECG-gated computed tomography (CT), which synchronizes image acquisition with the cardiac cycle to minimize motion artifacts and improve calcium scoring accuracy [3-5]. However, with advances in CT technology and improved temporal resolution, CAC scoring can also be performed on non-gated scans, with results that are often comparable to those obtained from gated studies [3]. Despite its clinical value, CAC scoring typically requires semi-manual annotation using dedicated software, which is time-consuming and subject to interobserver variability [6].

Recent advances in artificial intelligence (AI) and deep learning (DL) have enabled automated approaches to CAC detection and scoring. DL-based algorithms have been developed for use on both gated and non-gated CT scans, with the potential to streamline analysis and reduce reliance on manual interpretation [7-12]. Importantly, these tools have been validated in gated calcium scoring CT (CaCT) scans as well as in non-gated cohorts, including lung cancer screening populations, asymptomatic cardiac cohorts, and population-based screening studies, and have demonstrated prognostic value for future cardiovascular events [10, 13, 14]. Given the high prevalence of non-gated CT imaging in clinical practice and the growing interest in AI-based diagnostic tools, evaluating the performance of automated CAC assessment relative to standard manual scoring in both gated and non-gated CT cohorts is timely and clinically relevant.

A patient population that may benefit from automated CAC detection and risk stratification on non-gated CT is liver transplant candidates, in whom a high CAC burden is associated with increased risk of major adverse cardiac events after transplantation [15, 16].

Risk stratification by preoperative CAC scoring in liver transplant patients can help to predict postoperative complications [17]. In current practice, transplant programs often obtain non-gated chest CT scans to evaluate the lungs and mediastinum, followed by gated cardiac CT to assess for significant coronary artery disease. If CAC can be reliably assessed on the initial non-gated scan, the need for additional gated imaging may be reduced, thereby decreasing imaging burden, radiation exposure, and workflow complexity. More broadly, the ability of the deep-learning algorithm to effectively quantify CAC on non-gated CT scans may streamline diagnostic workflows and facilitate more widespread screening.

In this study, we evaluated the performance of a deep learning-based CAC algorithm (“DL-CAC”) for automatic identification and labeling of coronary calcium on both ECG-gated and non-gated non-contrast chest CT scans. The algorithm was previously evaluated at a single center using non-gated non-contrast thoracic CT scans [18]. Here, we report its performance in an independent, holdout dataset comprising a CaCT cohort, with standard-dose ECG-gated imaging and relatively thick slice reconstructions, and a lung cancer screening cohort, with low-dose non-gated imaging and variable slice thicknesses. In addition, we assessed agreement between manual CAC scoring on CaCT scans and automated CAC scoring on non-gated CT scans from an external cohort of liver transplant candidates.

## Methods

### Model Development

#### Training Cohort

The deep learning-based CAC inference algorithm (“DL-CAC”, 4D Medical Ltd., Woodland Hills, California, USA) was developed using 1260 anonymized, multi-center, retrospective, volumetric chest CT scans from the National Lung Cancer Screening Trial (NLST; https://cdas.cancer.gov/nlst), the Chronic Obstructive Pulmonary Disease Genetic Epidemiology (COPDGene) study (https://copdgene.org), and Multi-Ethnic Study of Atherosclerosis (MESA; https://mesa-nhlbi.org). These publicly available, de-identified, IRB-exempt datasets were divided into training and hyper-parameter tuning stages, with an ∼80:20 split ensuring balanced distribution across scanner vendors, reconstruction kernels, and slice thicknesses, as detailed in Table A1 of the appendix. This study was determined to be non human subjects research.

#### Deep-Learning Architecture

Model training for DL-CAC was conducted on an Amazon AWS g4dn.xlarge EC2 instance using the Swin UNETR architecture [19] within the MONAI framework. The algorithm first segments heart tissue and then classifies hyperintense calcifications within coronary arteries from a chest CT input. The training labels included: background = 0, right coronary artery (RCA) = 1, left main artery (LM) = 2, left anterior descending artery (LAD) = 3, left circumflex artery (LCx) = 4, and aorta = 5 (removed before final mask). Aortic calcifications were included to reduce false positives but were excluded from coronary calcium quantification. The input was a non-contrast volumetric chest CT image series that was preprocessed before DL model inference. Details on training settings and pre-processing are in the Appendix and Table A2.

The automated workflow outputs a calcium segmentation mask for each coronary artery, used to calculate quantitative measures including the Agatston score and arterial age, based on the McCollough et al. [20] method (mass calibration factor: 0.743 mg/(HU·cm^3^)).

#### Data Labeling, Augmentation, and Regularization Methods

Datasets containing ground truth CAC segmentation masks were produced by imaging technologists (3DR Labs; Louisville, Ky, USA) with at least three years of experience. Using ITK-SNAP software [21], technologists modified “high attenuation” segmentation masks generated by thresholding CT images and assigning pixel values ≥130 Hounsfield Units (HU) a value of 6. Labels were adjusted to classify RCA, LM, LAD, LCx, and ascending aorta calcifications, with the remaining label 6 values relabeled as “background.”

### Performance Testing Cohort #1 – Lung Cancer Screening and CaCTs

#### Cohort

All datasets used in the performance testing cohort were completely independent of model development and automatically analyzed with DL-CAC software. A total of 500 retrospective chest CT datasets met the input criteria from the following three databases: COPDGene, MESA, and Vega Imaging Informatics (https://vegainformatics.com, Ontario, CA, USA). Datasets included ECG-gated and non-gated, standard and low dose CT scans acquired on GE Medical, Siemens, Imatron, and Philips scanners. All datasets are de-identified and IRB-exempt. The cohort is fully described in Table A1 of the Appendix.

#### Inclusion/Exclusion Criteria

Exclusion criteria included the following: CTs from patients < 29 years old, contrast-enhanced CT scans, scans with significant respiratory motion, scans with severe metal artifacts, scans containing only non-thoracic anatomy (e.g., head CT). Inclusion criteria included pixel spacing ≤2.0 mm, slice spacing ≤ 3.0 mm, slice thickness ≤ 3.0 mm, complete field-of-view coverage of the heart, ECG-gated or non-gating acquisition, and either low dose or standard dose imaging.

#### Manual Labeling

The clinical gold standard for CAC scoring is the semi-automated identification of calcified regions within the coronary arteries by a qualified cardiac technologist in a chest CT scan. Ground truth annotations were obtained from qualified technologists from 3DR Labs. Three imaging technologists with at least three years’ experience used FDA-approved Vitrea cardiac software (Canon Medical Systems, CA, USA) to evaluate CT images, identifying pixels containing CAC and their location (i.e., RCA, LAD, LM, or LCx).

#### Risk classifications

A 5-category CVD risk classification was used with the following categories: none (Agatston score=0), minimal (1-10), mild (11-100), moderate (101-400), and extensive (>400) based on previously reported thresholds for Agatston scores in Rumberger et al. for calcium chest CT scans [22].

### Independent Testing Cohort #2 – Liver Transplant Cohort

#### Cohort

Non-gated chest CT and gated CaCT scans in patients who were evaluated for liver transplantation at the University of Chicago between July 9, 2012 - June 29, 2022, were examined. This cohort of non-gated chest CT scans is described in Table A1 of the appendix.

#### Inclusion/Exclusion Criteria

Inclusion criteria included age >18 years old, a non-gated non-contrast CT chest scan, and a CaCT scan acquired within one year of each other. A total of 129 subjects met these criteria. Four subjects were removed from analysis: two because calcium scores were missing from the electronic medical record and two because the input DICOM scans did not meet the minimum technical requirements for DL-CAC, resulting in a final total of 125 subjects.

#### Manual scoring

Vitrea software was used by a cardiac imaging fellow or attending physician (>3 years of experience) for manual segmentation and calcium scoring in major coronary arteries (i.e., LM, LAD, LCx, and RCA) on the CaCT scan only.

#### Risk classification

Cardiovascular risk was categorized as either low (≤10), moderate (11-400), or high (>400) based on liver transplant standards [23] and is used at University of Chicago. Low risk indicates no further cardiac testing was needed prior to transplantation. Moderate risk indicates further testing in the form of CT coronary angiography. High risk indicates a need for cardiac catheterization prior to liver transplantation.

### Statistical Analyses

DL-CAC software automatically analyzed all CT datasets. Reliability was assessed using Cohen’s κ or the intraclass correlation coefficient (ICC) [24] with 95% confidence interval (CIs) estimated with bootstrapping. Bias and limits of agreement (LoA) of Agatston scores were examined by Bland-Altman (BA) analysis. Mann-Whitney U-tests were used to assess differences between methods. Calculations were performed in Python using the scipy.stats library.

## Results

### Independent Testing Cohort #1 – Lung Cancer Screening and Calcium Scoring CTs

Analysis using the automated DL-CAC software was successful in 495/500 datasets. Ground truth Agatston scores were compared both overall and for each coronary artery, as well as by CVD risk classification. Five datasets were excluded due to incorrect heart segmentation. The average subject age was 64.3 ± 10.0 years old; 209 were male, 213 were female, and 73 did not have sex information available.

Expert-labeled Agatston scores for this cohort showed the following CVD risk distribution: none (n = 88), minimal (n = 69), mild (n = 124), moderate (n = 108), and extensive (n = 106). Agreement in CVD risk classification between ground truth and the DL-CAC software was strong, with Cohen’s κ = 0.936 (95% CI: [0.926 - 0.947]). Table 1 summarizes the reliability measurements for various subgroups within the cohort including ECG-gated, non-gated, low dose, and standard dose CT scans.

**Table 1.** Agreement, Cohen’s κ, for cardiovascular disease (CVD) risk classification across subgroups in the performance testing cohort #1 between ground truth and automated DL-CAC software.

| Groups | Subjects (N) | Cohen's $\kappa$ [95% CI] |
| --- | --- | --- |
| All | 495 | 0.936 [0.926 – 0.947] |
| Non-Gated | 245 | 0.907 [0.889 – 0.925] |
| ECG-Gated | 250 | 0.966 [0.956 – 0.977] |
| Standard Dose | 387 | 0.958 [0.948 – 0.967] |
| Low Dose | 108 | 0.861 [0.829 – 0.893] |

Agreement between Agatston scores from the DL-CAC software and ground truth was good for all coronary arteries (ICC > 0.79) and excellent overall (ICC = 0.947) (Table 2). Bland-Altman analysis indicated a bias ± LoA of −7.6 ± 405 Agatston units with larger errors at higher scores (Figure 1A). CVD risk classification discordance occurred in 70/495 (14%) subjects. The confusion matrix for DL-CAC CVD risk classification accuracy is shown in Figure 1B.

**Table 2.** ICC for Agatston score in total and in all four coronary arteries in performance testing cohort #1.

| Region | Subjects (N) | Agatston ICC [95% CI] | Bias [95% CI] | Limits of Agreement (LoA) |
| --- | --- | --- | --- | --- |
| Total | 495 | 0.947 [0.94 – 0.96] | -7.6 [-25.8 – 10.7] | -412 – 397 |
| LAD | 495 | 0.896 [0.88 – 0.91] | -3.6 [-13.7 – 6.4] | -226 – 219 |
| LM | 495 | 0.793 [0.76 – 0.82] | -5.2 [-8.3 – -2.1] | -73 – 63 |
| LCx | 495 | 0.879 [0.86 – 0.90] | 10.8 [3.0 – 18.5] | -162 – 183 |
| RCA | 495 | 0.862 [0.84 – 0.88] | -9.5 [-21.9 – 2.9] | -285 – 266 |

**Figure 1.**
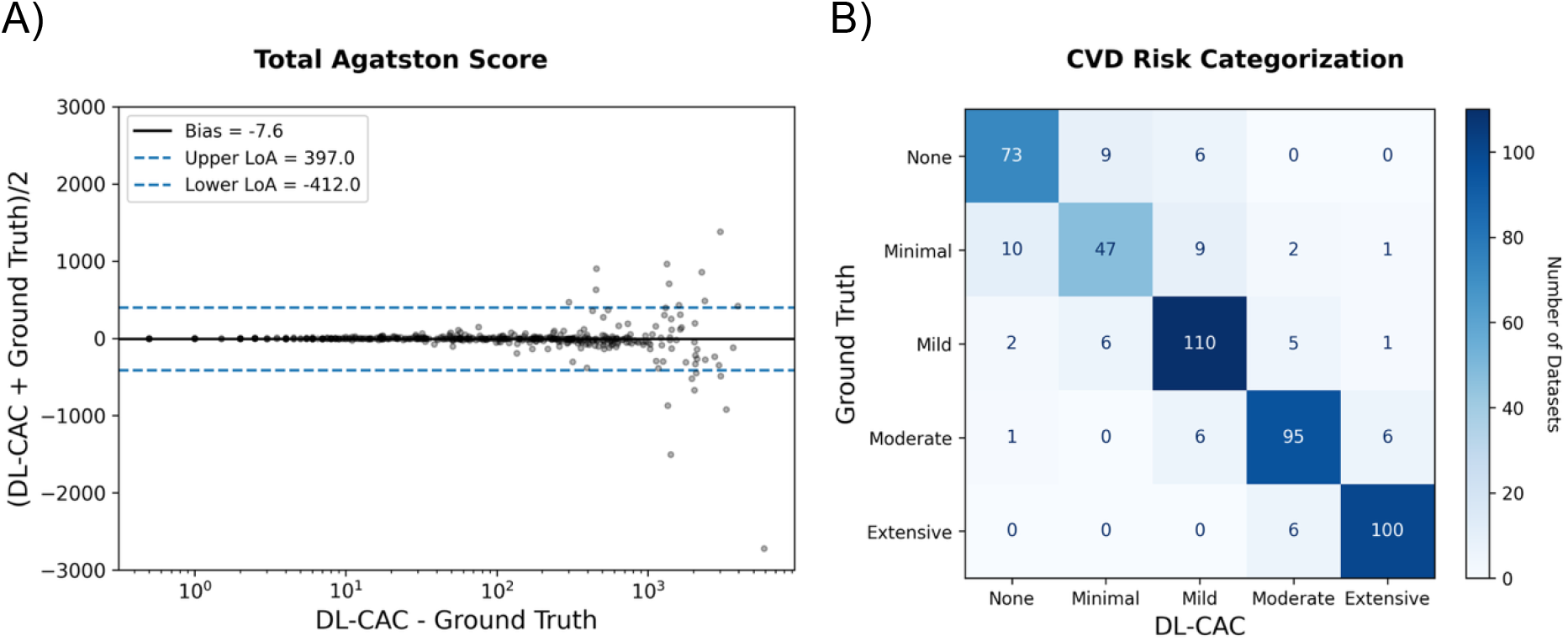
Performance testing cohort #1. (**A**) Bland-Altman plot of the total Agatston scores between the DL-CAC algorithm and expert-labeled ground truth. (**B**) Confusion matrix showing the agreement between none (Agatston score = 0), minimal (1-10), mild (11-100), moderate (101-400), and extensive (>400) cardiovascular disease (CVD) risk classification.

### Independent Testing Cohort #2 – Liver Transplant Candidate Cohort

In the liver transplant cohort, the DL-CAC algorithm’s Agatston scores on non-gated CTs were compared with expert measurements on ECG-gated CaCTs using Bland-Altman analysis (Figure 2A). CVD risk classification was compared via a confusion matrix between the AI algorithm and manual scoring (Figure 2B).

**Figure 2.**
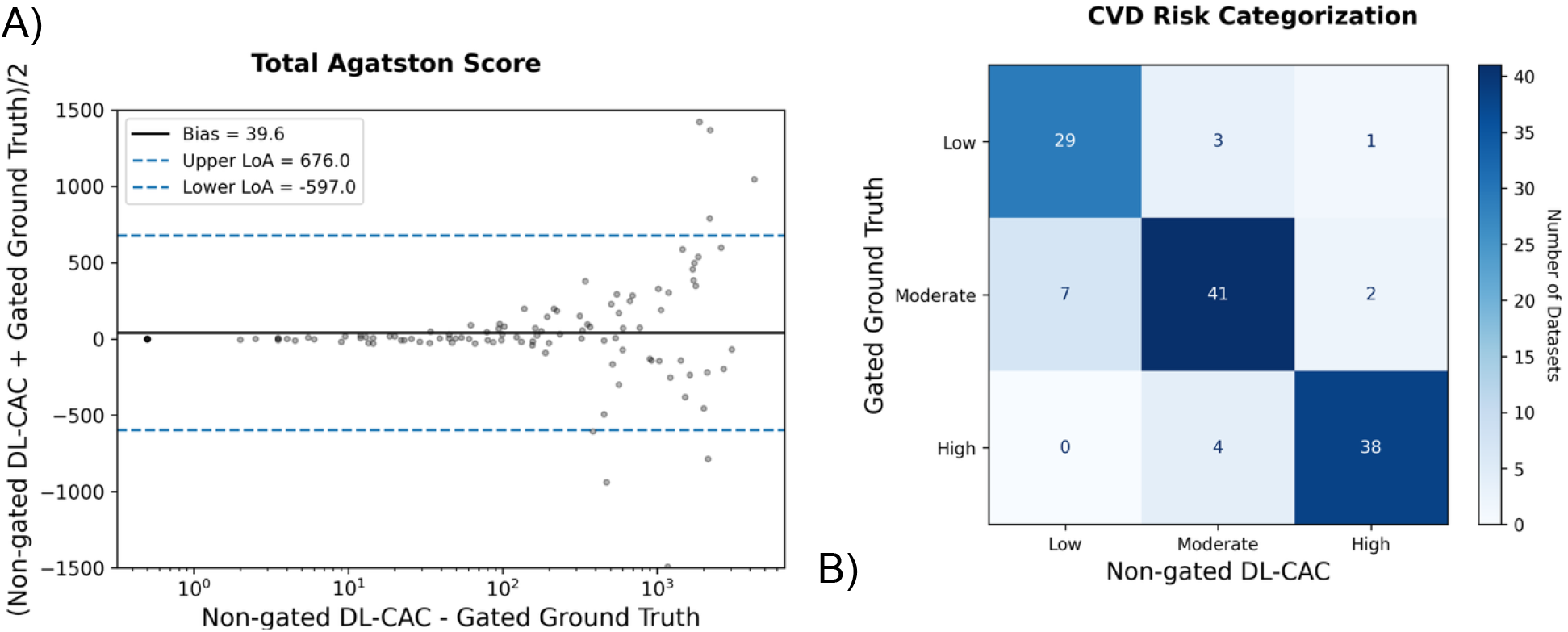
Performance testing cohort #2. (**A**) Bland-Altman plot illustrating differences between total Agatston scores measured using the DL-CAC algorithm’s scoring on non-gated scans versus expert manual ground truth scoring on ECG-gated scans. (**B**). Confusion matrix showing the agreement between low (Agatston score = 0-10), moderate (11-400), and high (>400) cardiovascular disease (CVD) risk classification.

Cohen’s κ was 0.79 indicated moderate agreement and the Mann-Whitney U-test (p-value = 0.752) showed no significant difference in risk levels assigned by DL-CAC on non-gated scans versus expert scoring on gated scans (Table 3). For diagnostic performance on moderate/high-risk cases, DL-CAC had a sensitivity of 93.4% and a specificity of 88.2%. For high-risk cases only, DL-CAC had a sensitivity of 90.4% and a specificity of 96.4% (Table 3).

**Table 3.** (Top) Concordance performance between DL-CAC software and ground truth labels including the agreement for 3-category cardiovascular disease (CVD) risk classification and Bland-Altman (BA) analysis. (Bottom) DL-CAC software detection performance in liver transplant candidates with moderate-to-high CVD risk. Moderate CVD risk = 11-400 Agatston score, and high CVD Risk = >400 Agatston score.

| Concordance Performance |  |  |
| --- | --- | --- |
|  | Metric | Value |
| Agreement for CVD Risk | Cohen's $\kappa$ | 0.79 |
|  | Mann-Whitney U-test | 7642, p=0.752 |
| Bland Altman for Total Agatston | BA Bias (ground truth – DL-CAC) | 39.6 |
|  | BA Limits of agreement | 636.5 |

| Detection Performance |  |  |
| --- | --- | --- |
| CVD Risk | Moderate + High | High |
| Sensitivity | 93.4% | 90.4% |
| Specificity | 88.2% | 96.4% |

CVD risk discordance was observed in 17/125 (13.6%) subjects. Non-gated scans often had lower Agatston scores because motion blurred small calcium lesions, causing missed or miscounted calcifications. In one case, motion blurring increased apparent lesion size. Examples are shown in Figure 3.

**Figure 3.**
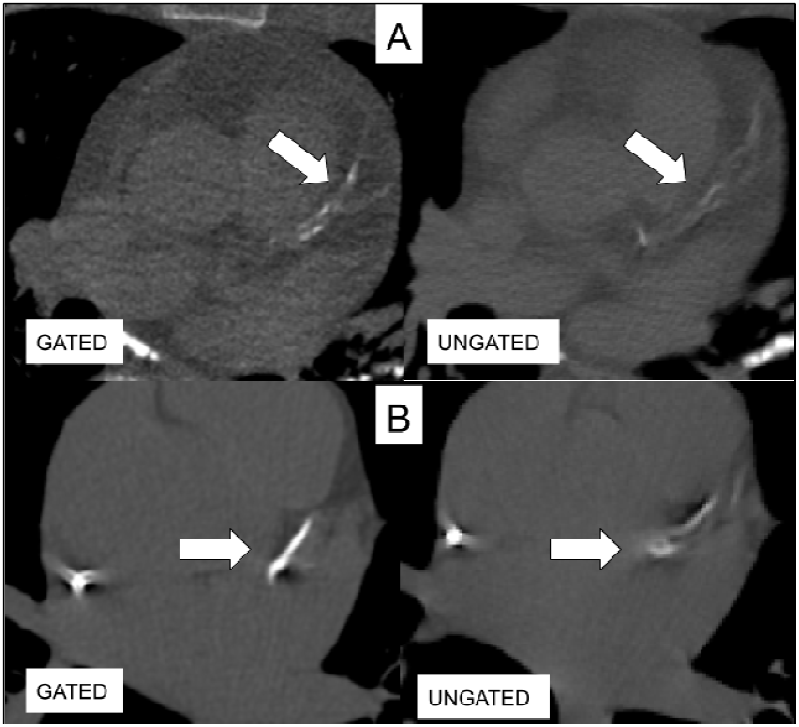
(**A**). Blurred calcium in the non-gated scan causing underestimation of Agatston score. (**B**). Motion causing the calcium lesion to be classified as artifact, leading to Agatston score underestimation.

## Discussion

We developed and evaluated an automated deep-learning algorithm for CAC scoring in two independent testing cohorts. First, we tested DL-CAC on a pooled cohort consisting of non-gated thoracic CT scans and traditional gated CAC CT scans. Results demonstrated excellent agreement between the algorithm and expert-derived ground truth CAC labels for both CVD risk classification (κ = 0.936 [0.926 – 0.947]) and total Agatston score (ICC = 0.947 [0.94 – 0.96]). In comparison, alternative DL-based CAC algorithms that perform a 3-category CVD risk classification, demonstrated a comparable κ = 0.925 [0.897 – 0.955] [25], and κ = 0.870 [0.850 – 0.890] [14]. Our algorithm exhibited minimal bias and acceptable limits of agreement when compared to ground truth. It also performed consistently across subgroups, including low dose, standard dose, ECG-gated, and non-gated scans.

Notably, the lowest agreement was observed in low-dose scans which consisted primarily of non-gated scans (102/108 scans). Noise, patient motion, and imaging protocol are known confounders that can degrade CAC quantification performance. For example, Suh et al. demonstrated in 452 subjects with ECG-gated cardiac CTs and low dose CTs that automated CAC analysis showed very high agreement for CVD risk classification (κ = 0.918-0.972) on ECG-gated CaCTs yet heterogeneous agreement (κ = 0.748-0.924) on low dose CTs depending on the institution (i.e., data source) [26].

Second, we compared algorithm performance in non-gated CT scans with manual scoring from gated scans within a liver transplant candidate population from a tertiary imaging center. The substantial concordance (κ = 0.79) observed between the deep-learning algorithm in non-gated scans and manual scoring on gated scans highlights the potential of AI-driven tools to accurately assess coronary artery calcification without the need for gating. In particular, the high diagnostic performance to identify moderate+high risk cases (sensitivity/ specificity = 93.4%/88.2%) or high-risk cases only (sensitivity/specificity = 90.4%/96.4%) could help identify which liver transplant candidates are at risk of major adverse cardiac events and postoperative complications [15].

In clinical practice, using a single CT scan instead of the conventional approach of acquiring both a chest CT and a gated CaCT could reduce radiation exposure and cost and would offer substantial benefit to patients. Moreover, non-gated CT scans are more easily acquired and can be performed more rapidly compared to gated scans, which require synchronization with the cardiac cycle and may involve additional patient preparation. Given the larger population of patients imaged with chest CT for indications other than for assessment of coronary artery calcium, this algorithm could also be used for population screening to identify patients without a history of coronary artery disease who are at high risk for an acute cardiovascular event in the near future.

The ability of the deep-learning algorithm to effectively quantify CAC on non-gated CT scans may streamline diagnostic workflows and facilitate more widespread screening. For liver transplant candidates, reducing the reliance on gated CT could alleviate some of the burdens associated with diagnostic imaging and improve overall efficiency in clinical settings. The value of risk stratification by CAC scoring in liver transplant subjects has been shown elsewhere [17, 26, 27]. It is not uncommon for transplant programs to screen for underlying lung and mediastinal disease using chest CT and then acquire a gated cardiac CT to rule out significant coronary artery disease. The DL-CAC algorithm effectively assessed the CVD risk directly from chest CTs originally acquired for non-cardiac purposes, eliminating the need for the additional and possibly redundant cardiac-gated CT.

In conclusion, our study indicates that deep learning-based automated algorithms can effectively analyze non-gated CT scans as an alternative to gated CT for calcium scoring. The observed agreement with traditional methods suggests that AI analysis on non-gated CTs could potentially replace the need for gated CTs for CAC quantification, optimizing imaging practices. Future research should validate these findings further and explore integrating AI-driven tools into routine clinical practice.

## Data Availability

All data produced in the present study are available upon reasonable request to the authors.

## Appendix

**Table A1.** Data characteristics of the CT parameters used in the model’s development and testing.

|  | **Model Development** |  | **Model Testing** |  |
| --- | --- | --- | --- | --- |
|  | **Training** | **Tuning** | **Performance Cohort #1** | **Performance Cohort #2** |
| **Datasets Total (N):** | 1000 | 260 | 495 | 129 |
| **Database (N)** |  |  |  |  |
| NLST | 219 | 64 | 0 | 0 |
| COPDGene | 392 | 94 | 68 | 0 |
| MESA | 389 | 102 | 83 | 0 |
| Vega Imaging | 0 | 0 | 344 | 0 |
| University of Chicago | 0 | 0 | 0 | 129 |
| **CT Manufacturer (N)** |  |  |  |  |
| GE | 214 | 48 | 324 | 99 |
| Philips | 10 | 4 | 1 | 30 |
| Siemens | 457 | 121 | 94 | 0 |
| Imatron | 301 | 6 | 76 | 0 |
| Toshiba/Canon | 0 | 0 | 0 | 0 |
| **Gating (N)** |  |  |  |  |
| ECG-Gated | 389 | 102 | 250 | 0 |
| Non-Gated | 611 | 158 | 245 | 129 |
| **Slice Thickness (mm)** |  |  |  |  |
| $\leq 1.0$ | 0 | 0 | 68 | 126 |
| $1.0 < x \leq 2.0$ | 494 | 115 | 177 | 3 |
| $2.0 < x \leq 3.0$ | 59 | 15 | 249 | 0 |
| $> 3.0$ | 26 | 12 | 1 | 0 |
| **Dosage** |  |  |  |  |
| Low Dose | 805 | 215 | 108 | 13 |
| Standard Dose | 177 | 36 | 387 | 116 |
| N/A | 18 | 9 | 0 | 0 |
| **Pixel Size (mm)** |  |  |  |  |
| $\leq 0.6$ | 106 | 28 | 171 | 8 |
| $0.6 < x \leq 0.8$ | 885 | 227 | 205 | 109 |
| $0.8 < x \leq 1.0$ | 9 | 5 | 119 | 12 |
| $> 1.0$ | 0 | 0 | 0 | 0 |
| **CT Reconstruction Kernels** |  |  |  |  |
| B | 10 | 4 | 1 | 29 |
| B30F | 124 | 34 | 7 | 0 |
| B31F | 125 | 36 | 20 | 0 |
| B35f | 2 | 1 | 4 | 0 |
| B40s | 0 | 0 | 52 | 0 |
| B45F | 28 | 4 | 0 | 0 |
| B46F | 9 | 4 | 0 | 0 |
| B50F | 42 | 13 | 0 | 0 |
| B80F | 43 | 8 | 0 | 0 |
| BF40D* | 51 | 11 | 0 | 0 |
| BONE | 48 | 13 | 0 | 0 |
| D | 0 | 0 | 0 | 1 |
| I31F* | 26 | 8 | 8 | 0 |
| I44F | 3 | 0 | 0 | 0 |
| LUNG | 1 | 1 | 0 | 3 |
| QR44D* | 6 | 2 | 0 | 0 |
| Sa36d | 0 | 0 | 3 | 0 |
| SHARP | 5 | 6 | 0 | 0 |
| SHARP R62 | 0 | 0 | 5 | 0 |
| STANDARD | 165 | 34 | 324 | 96 |
| N/A | 0 | 0 | 71 | 0 |

**Table A2.** DL-CAC model training settings.

|  |  |
| --- | --- |
| Epochs | 300 |
| Batch size | 5 |
| Optimizer | AdamW |
| Learning rate | 0.001 |
| Loss function | Dice Focal Loss |

## Data augmentation

During training, the images were augmented using the following: 3D affine transformations (random ± 5° rotations in all planes, random ± 5% shear in all planes) and random crops around the region of interest (heart segmentation).

## Pre-processing

During inference, the input CT image is cropped to a bounding box of the heart, resized to a cube with edge length of 224 pixels, and pixel intensity normalized to the 0 to 1 range using - 300 to 300 Hounsfield units as the lower and upper boundaries. The 224-pixel length cube is decomposed into smaller cubes with an edge length of 64 pixels, with a stride that overlaps 25% between cubes. Inference is performed on each 64-pixel cube, and results are averaged and stitched back together to fill out the original 224-pixel cube. Final output is resized to the original CT input image and post processed to remove the non-CAC labels (aorta = = 5). Segmented pixels that do not mask CT pixels with Hounsfield unit values greater or equal to 130 HU are removed.

## Notes

**Conflict of Interest:** Kai D. Ludwig, Charles R. Hatt, and Lauren Keith: Employed by 4D Medical Ltd. Amit R. Patel: Has research grants/support from GE Healthcare, CircleCVI, Area19, Neosoft, and Siemens Healthineers. Jonathan H. Chung: Consultant for Boehringer Ingelheim, Imbio/4D Medical, AstraZeneca, and Daiichi Sankyo, speaker for Boehringer Ingelheim, and clinical advisor for Riverain.

### Competing Interest Statement

Kai D. Ludwig, Charles R. Hatt, and Lauren Keith: Employed by 4D Medical Ltd. Amit R. Patel: Has research grants/support from GE Healthcare, CircleCVI, Area19, Neosoft, and Siemens Healthineers. Jonathan H. Chung: Consultant for Boehringer Ingelheim, Imbio/4D Medical, AstraZeneca, and Daiichi Sankyo, speaker for Boehringer Ingelheim, and clinical advisor for Riverain.

### Funding Statement

This study was funded by 4D Medical Ltd.

